# Life stage-specific effects of genetic susceptibility to higher body size on body fat and lean mass: prospective cohort study

**DOI:** 10.1101/2022.04.28.22274413

**Authors:** Scott Waterfield, Tom G. Richardson, George Davey Smith, Linda M. O’Keeffe, Joshua A. Bell

## Abstract

**Background/Objectives:** Separate genetic variants are associated with larger body size in childhood and adulthood. Whether these variants predominantly influence adiposity, and whether these truly differ by life stage is unknown. We examined how genetic variants influence total body fat and total lean mass trajectories from childhood to young adulthood.

**Methods:** Data were from the Avon Longitudinal Study of Parents and Children birth cohort. Sex-specific genetic risk scores (GRS) for childhood and adulthood body size were generated, and dual-energy X-ray absorptiometry scans were used to measure total body fat and lean mass 6 times between ages 9-25y. We used mutually-adjusted multilevel linear spline models to examine the independent sex-specific associations of childhood and adulthood body size GRSs with fat and lean mass trajectories from 9 -25y.

**Results:** In males, the childhood and adulthood GRS were associated with similar differences in fat mass from 9 to 18y; 8.3% (95% confidence interval (CI): 5.1,11.6)) and 7.5% (95% CI: 4.3,10.8) higher fat mass at 18y per standard deviation (SD) higher childhood and adulthood GRS respectively. At 25y, childhood GRS associations with fat mass attenuated while adulthood GRS associations remained similar to those at 18y for males. Among females, associations for the childhood GRS were almost two-fold stronger than the adulthood GRS from 9 to 18y: 10.5% (95% CI: 8.5, 12.4) higher fat mass at 9y per SD higher childhood GRS compared with 5.1% (95% CI 3.2, 6.9) per SD higher adulthood GRS. At 25y, associations of the childhood and adulthood GRS with fat mass were similar; 5.0% (95% CI: 2.5, 7.5) and 5.8% (95% CI: 3.3, 8.3) higher fat mass per SD higher childhood and adulthood GRS respectively: Lean mass effect sizes were much smaller.

**Conclusions:** Genetic variants for body size are more strongly associated with adiposity than lean mass from childhood to early adulthood; childhood variants are more strongly associated with adiposity in females until early adulthood whereas childhood and adulthood variants are similarly associated with adiposity across early life in males. Findings may inform selection of instruments for life stage-specific adiposity in future Mendelian randomization studies.

## Introduction

Excess adiposity is now a global pandemic, with 64% of UK adults being overweight or obese, and 34% of UK children being overweight or obese by age 10 years^1^. Overweight and obesity are associated with negative health outcomes, such as accelerated biological aging (as measured by epigenetic age analysis)^2,3^, and a number of life-limiting diseases such as cardiovascular disease and cancer ^4–7^. A host of environmental factors such as socioeconomic disadvantage^8^, energy dense diets, and physical inactivity^9^ likely influence the population rates of excessive adiposity.

Genome-wide association studies (GWAS) have established that individual susceptibility to higher adiposity is influenced in part by genetic variation, with hundreds of single nucleotide polymorphisms (SNPs) now found to be robustly associated with adiposity as measured indirectly using body mass index (BMI)^10^. Most of these genetic studies have focused on BMI measured in adulthood, the largest of which are meta-analyses of data from the Genetic Investigation of Anthropometric Traits (GIANT) consortium and the UK Biobank^11^. Genetic variants associated with BMI measured in childhood were first discovered by the Early Growth Genetics (EGG) consortium^12^, and have recently been expanded upon using self-recalled childhood body size relative to peers from the larger UK Biobank dataset. These latter findings suggest that large numbers of different SNPs are associated with childhood body size, with only moderate overlap with adulthood body size SNPs^13^. However, due to the lack of objective measures of body composition in the aforementioned studies, it is still unclear whether these SNPs predominantly raise adiposity as opposed to lean mass.

Furthermore, most studies have not had repeat assessments of objective body composition measures such as fat and lean mass available across different life stages and thus, whether the influence of different body size SNPs is truly confined to specific periods of the life course is not well understood. By examining associations of body size SNPs with objective measures of body composition, taken repeatedly across different life stages, insights into the time-sensitive effects of these SNPs on adiposity or lean mass would be enabled. This would aid the interpretation of Mendelian randomization (MR) studies which use these SNPs as instruments to study the impact of life-stage adiposity on health outcomes.

In this study, we examined the associations of SNPs for childhood and adulthood body size using genetic risk scores (GRSs) with trajectories of dual-energy x-ray absorptiometry (DXA) total, trunk, and peripheral (arms + legs) fat mass, and total lean mass from 9 to 25 years (y) in the Avon Longitudinal Study of Parents and Children (ALSPAC) birth cohort. Analyses were performed separately by sex using sex-specific GRSs to examine how genetic influences on fat and lean mass at different life stages may differ between males and females.

## Methods

### Study participants

ALSPAC is a prospective birth cohort study in south west England. Pregnant women resident in one of the three Bristol-based health districts with an expected delivery date between April 1, 1991 and December 31, 1992 were invited to participate. The study has been described elsewhere in detail and ethical approval for the study was obtained from the ALSPAC Ethics and Law Committee and the Local Research Ethics Committees^14–16^.

ALSPAC initially enrolled a cohort of 14,541 pregnancies, from which 14,062 live births occurred, with 13,998 alive at 1 year. Follow-up has included parent and child completed questionnaires, links to routine data and clinic attendance. The present analyses were restricted to offspring participants who were first-born and of a white ethnicity given that the source GWAS for body size was based on individuals of European ancestry.

Research clinics were held when these offspring participants were approximately 9, 10, 11, 13, 15, 18, and 25 years old. Data for 25 years of age were collected and managed using REDCap electronic data capture tools hosted at the University of Bristol. REDCap (Research Electronic Data Capture) is a secure, web-based software platform designed to support data capture for research studies^17^. The study website contains details of all the data that is available through a fully searchable data dictionary http://www.bristol.ac.uk/alspac/researchers/access/.

### Childhood and adulthood body size GRSs

Separate sets of SNPs for childhood body size and adulthood body size were used for males and females based on sex-specific GWAS; 134 childhood female, 212 adulthood female, 69 childhood male, and 158 adulthood male SNPs^13^. GRSs were constructed using PLINK 1.9, with effect alleles and beta coefficients from the GWAS used as external weightings.

Standard scoring was applied by multiplying the effect allele count (or probabilities if imputed) at each SNP (values 0, 1, or 2) by its weighting, summing these, and dividing by the total number of SNPs used. Each score therefore reflects the average per-SNP effect on increasing a category of childhood or adulthood body size (separately).

### Body fat and lean mass measures

Total body fat (less head), central (trunk) fat, peripheral (legs + arms) fat, and total lean mass (less head), each in kg, were measured using whole body DXA scans performed at the ALSPAC clinics undertaken at ages 9, 11, 13, 15, 18, and 25 using a Lunar prodigy narrow fan beam densitometer. Scans were screened for anomalies, motion, and material artefacts, and realigned when necessary.

### Associations of childhood and adulthood body size GRSs with trajectories of body fat and lean mass

In males and females separately, we used multilevel linear spline models to examine associations of a standard deviation (SD) higher childhood and adulthood body size GRS with trajectories of DXA total, trunk, and peripheral fat mass, and total lean mass, from 9y to 25y. As our goal was to examine independent associations of the childhood and adulthood GRS with changes over time in body composition, all analyses were performed with mutual adjustment for GRSs, i.e., the childhood GRS model was adjusted for the adulthood GRS and vice versa. Multilevel models estimate the mean trajectory of the outcome while accounting for the non-independence (i.e., clustering) of repeated measurements within individuals, change in scale and variance of measures over time, and differences in the number and timing of measurements between individuals (using all available data from all eligible participants under a Missing at Random (MAR) assumption)^18^. Linear splines allow knot points to be fit at different ages to derive periods in which change is approximately linear. Knot points were fitted at age 13 and 15 as per previous work^19,20^, with an additional knot point added at 18 for the extension of trajectories to 25 years. These linear spline multilevel models included two levels: measurement occasion and individual. All DXA models included sex-and age-specific adjustments for height as previously described^21^. Inclusion criteria for analyses were as follows: availability of sex-specific GRS data for both childhood and adulthood, and at least one DXA measure for the specific outcome being analysed.

To aid interpretation, GRSs were internally standardised by centring around the sample mean and dividing by the SD: (individual GRS minus mean(GRS) divided by SD(GRS). Fat-based DXA measures had skewed distributions on most occasions and were therefore log transformed before modelling took place. To aid comparability with fat-based measures, lean mass was also log transformed. The sex-specific effect of each GRS (per SD) on each outcome trajectory was then estimated by including an interaction term between the standardised GRS and the intercept (value of the outcome at 9 years) and each linear spline period, providing estimates for the association between an SD increase in each GRS and the intercept and each linear spline period. Following analysis, these estimates were then used to calculate the mean predicted trajectory of fat and lean mass for individuals with an SD higher (relative to the mean) childhood and adulthood GRS. Note that for outcomes which were log transformed these graphs are displayed in original units by back transforming from the log scale. The difference in fat and lean mass at different ages from 9 to 25 years per SD higher childhood and adulthood GRS was also calculated. Differences and confidence intervals were calculated on the log-scale, then back-transformed to a ratio of geometric means and converted to and displayed as percentage differences in predicted fat and lean mass at different ages from 9 to 25 years per SD higher childhood and adulthood GRS. Sensitivity analysis models as described above were also run using a a) non-sex specific GRS, b) without mutual adjustment and c) non sex-specific GRS without mutual adjustment. All trajectories were modelled in MLwiN (V3.05)^22^, called from R statistical software using the R2MLwiN library^23^.

This study is reported in line with STROBE guidelines (see **Supplemental item 1**).

## Results

### Sample characteristics

A total of 6926 individuals (3511 females and 3415 males) were included in our analyses. **Table 1** describes several socio-demographic characteristics of participants included in our analyses. Males and females included in analyses did not differ based on their parental socioeconomic backgrounds. Females had lower birthweight, younger ages at onset of puberty and higher fat mass at ages 9y and 25y than males. Males had higher lean mass at 9 and 25y than females. **Supplementary Table 1** compares characteristics of participants included in analyses vs. those excluded from analyses due to missing data (either not having genetic data and/or at least one DXA measure). Participants included in analyses had a higher proportion of white ethnicity, had parents who had higher educational attainment and household social class, and mothers who were less likely to have smoked during pregnancy than those who were excluded from analyses.

**Table 1.** Characteristics of ALSPAC participants included in analyses

|  | **Females**<br>**N = 3511**<br>**n (%)** | **Males**<br>**N = 3415**<br>**n (%)** |
| --- | --- | --- |
| **Household social class\*** |  |  |
| Professional | 466 (15.1) | 476 (15.9) |
| Managerial & Technical | 1395 (45.3) | 1332 (44.4) |
| Non-Manual | 748 (24.3) | 745 (24.8) |
| Manual | 321 (10.4) | 326 (10.9) |
| Part skilled & unskilled | 148 (4.8) | 121 (4.0) |
| **Mother's highest educational qualification†** |  |  |
| Less than O level | 706 (22.0) | 729 (23.1) |
| O level | 1148 (35.8) | 1121 (35.6) |
| A level | 828 (25.9) | 807 (25.6) |
| Degree or above | 521 (16.3) | 494 (15.7) |
| **Mother's partner's highest educational qualification†** |  |  |
| Less than O level | 931 (29.7) | 831 (27.2) |
| O level | 675 (21.5) | 683 (22.3) |
| A level | 872 (27.8) | 855 (27.9) |
| Degree or above | 658 (20.9) | 690 (22.6) |
| **Maternal smoking during pregnancy (first three months)‡** | 606 (18.6) | 623 (19.5) |
|  | **Mean (SD)** | **Mean (SD)** |
| **Birth weight (kg)** | 3.39 (0.48) | 3.51 (0.54) |
| **Puberty Timing: Age at peak height velocity (years)‡** | 11.75 (0.82) | 13.58 (0.92) |
| **Body composition measures** |  |  |
| DXA total fat mass age 9 clinic (kg) | 9.7 (5.0) | 7.3 (4.8) |
| DXA total fat mass age 25 clinic (kg) | 24.8 (10.1) | 20.7 (9.7) |
| DXA trunk fat mass age 9 clinic (kg) | 3.9 (2.5) | 2.9 (2.3) |
| DXA trunk fat mass age 25 clinic (kg) | 11.5 (6.1) | 10.6 (5.9) |
| DXA peripheral fat mass age 9 clinic (kg) | 5.2 (2.4) | 3.9 (2.4) |
| DXA peripheral fat mass age 25 clinic (kg) | 12.4 (4.9) | 9.2 (3.9) |
| DXA total lean mass age 9 clinic (kg) | 23.6 (3.2) | 25.5 (3.0) |
| DXA total lean mass age 25 clinic (kg) | 41.2 (5.3) | 57.1 (7.5) |

**Legend:** The total N for these characteristics differs somewhat from sample sizes used in analyses as these complete data on these characteristics was not required for our analyses.
DXA - dual-energy X-ray absorptiometry.
\*Household social class was measured as the highest of the mother's or her partner's occupational social class using data on job title and details of occupation collected about the mother and her partner from the mother's questionnaire at 32 weeks gestation. Social class was derived using the standard occupational classification (SOC) codes developed by the United Kingdom Office of Population Census and Surveys and classified as I-professional, II-managerial and technical, IIINM-non-manual, IIIM-manual, and IV&V part skilled occupations and unskilled occupations.
†Education status was recorded via a questionnaire of the mother, asking for her highest level of education, and her partner's highest level of education at 32 weeks of gestation. Responses are categorised into less than O-level (including vocational courses and the certificate of secondary education (CSE), O-level (taken at 16 years), A-levels (taken at 18 years), or university degree (or higher).
‡Maternal smoking in the first trimester was self-reported via questionnaire of the mothers; form of smoking was defined as: cigarettes, cigars, pipe, or 'other', all of which we combine into a binarized value of having smoked or not having smoked during pregnancy.
‡Puberty timing was estimated via age at peak height velocity based on the Superimposition by Translation And Rotation (SITAR) methodology<sup>24</sup>.

### Estimated effects of childhood and adulthood body size GRSs on trajectories of fat mass

**Supplementary Figure 1A-B** shows the mean sex-specific trajectories of total fat mass from 9-25y for 1) participants with the mean childhood GRS and mean adulthood GRS, 2) participants with a 1 SD higher childhood GRS compared with the mean and 3) participants with a 1 SD higher adulthood GRS compared with the mean.

Compared with a male with the mean childhood and adulthood GRS, a 1 SD higher childhood GRS or adult GRS was associated with 8.8% (95% Confidence interval (CI) 6.3, 11.3) and 6.1% (95% CI 3.7, 8.6) higher total fat mass at 9y, respectively, and similar rates of change in total fat mass from 9 to 25y among males (**Figure 1A** and **Table 2**). This resulted in the persistence of associations of childhood and adulthood GRSs with total fat mass at 9y up to age 18y; associations for both GRSs were of comparable magnitude at each age and were broadly similar across these ages. For example, associations of a 1 SD higher childhood and adulthood GRS with total fat mass were similar at 18y with 8.3% (95% CI 5.1, 11.6) and 7.5% (95% CI 4.3, 10.8) higher total fat mass at 18y per SD higher childhood and adulthood GRS, respectively. At age 25y, associations for the childhood GRS attenuated while associations for adulthood GRS persisted (difference per SD higher childhood GRS: 2.9% (95% CI: -1.0, 6.9), difference per SD higher adulthood GRS: 5.3% (95% CI: 1.3, 9.5)).

**Figure 1.**
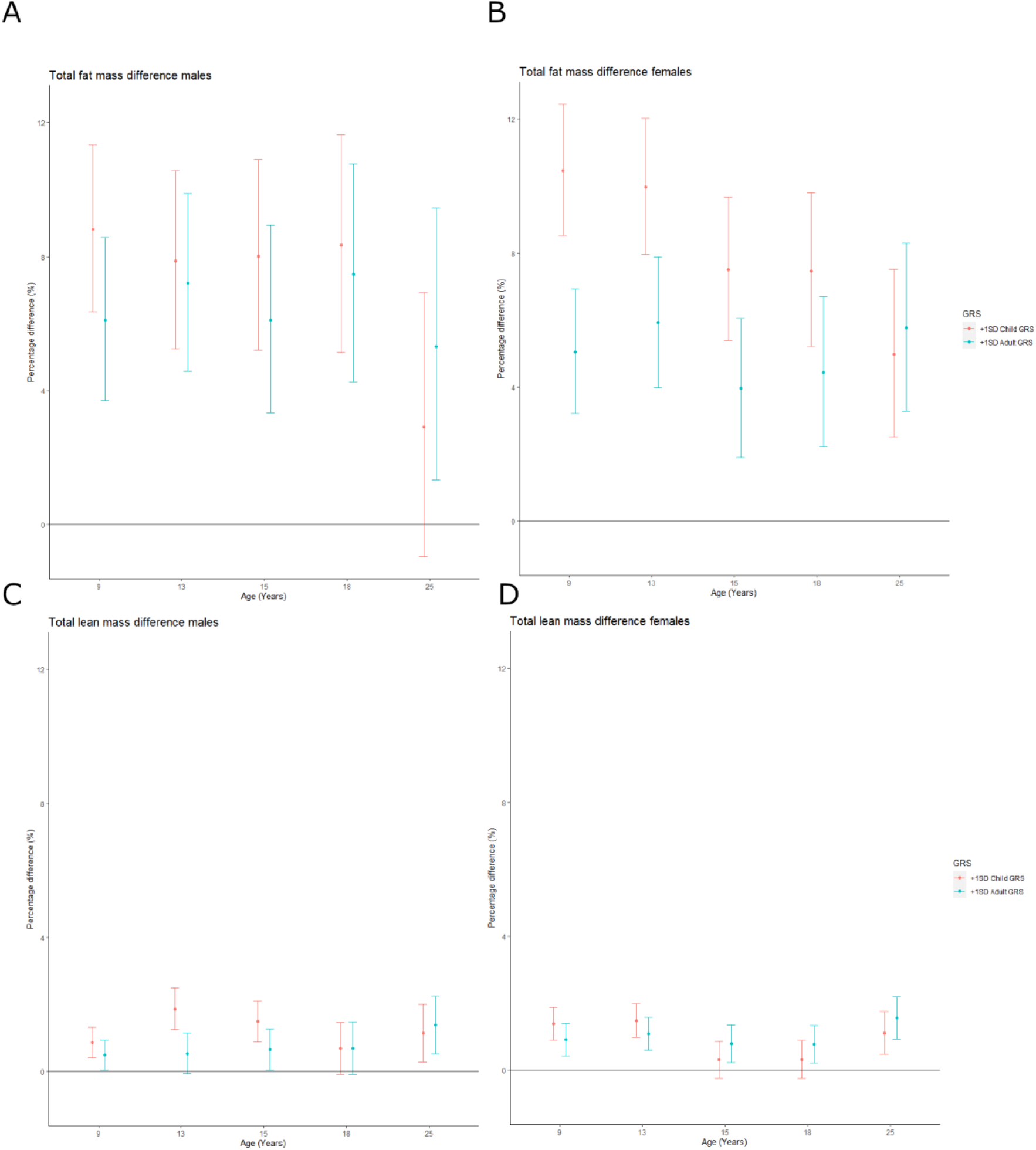
Mean difference in fat (A, B) and lean mass (C, D) in females and males from 9 to 25 years by child and adult GRS. Differences shown are for a participant with a 1 SD higher childhood GRS and a 1 SD higher adulthood GRS compared with the mean (participant with the mean childhood and adulthood GRS). Differences are derived from models which include mutual adjustment for each score. Mean trajectory is centred on the sex-specific mean of the child (male mean: 0.0098 (SD:0.0008), female mean: 0.0086 (SD: 0.0005)) and adult (male mean: 0.0076 (SD:0.0004), female mean: 0.0079 (SD: 0.0004)) GRS. The difference in fat mass per SD higher child or adult GRS is back transformed from the log scale for ease of interpretation and is a ratio of geometric means, expressed as a percentage difference.

**Table 2.** Mean trajectory and mean difference in trajectory of total fat mass from 9 to 25y per SD higher child GRS and adult GRS in males, with mutual adjustment for each score

| Age (years) | Mean trajectory (95% CI)* | Age (years) | Mean difference (95% CI) per SD higher child GRS | Mean difference (95% CI) per SD higher adult GRS |
| --- | --- | --- | --- | --- |
| 9 (kg) * | 5.6 (5.4, 5.8) | 9 (% difference) ‡ | 8.8 (6.3, 11.3) | 6.1 (3.7, 8.6) |
| 9-13 (%/y) † | 15.3 (14.2, 16.4) | 9-13 (% difference/y) § | -0.2 (-0.8, 0.3) | 0.3 (-0.3, 0.8) |
| 13 (kg) * | 9.9 (9.7, 10.2) | 13 (% difference) ‡ | 7.9 (5.2, 10.6) | 7.2 (4.6, 9.9) |
| 13-15 (%/y) † | -6.9 (-7.8, -5.9) | 13-15 (% difference/y) § | 0.1 (-0.9, 1.1) | -0.5 (-1.5, 0.5) |
| 15 (kg) * | 8.6 (8.4, 8.8) | 15 (% difference) ‡ | 8.0 (5.2, 10.9) | 6.1 (3.3, 8.9) |
| 15-18 (%/y) † | 10.6 (9.8, 11.4) | 15-18 (% difference/y) § | 0.1 (-0.7, 0.9) | 0.4 (-0.4, 1.2) |
| 18 (kg) * | 11.6 (11.3, 12.0) | 18 (% difference) ‡ | 8.3 (5.1, 11.6) | 7.5 (4.3, 10.8) |
| 18-25 (%/y) † | 7.9 (7.4, 8.3) | 18-25 (% difference/y) § | -0.7 (-1.2, -0.3) | -0.3 (-0.7, 0.2) |
| 25 (kg) * | 19.8 (19.1, 20.5) | 25 (% difference) ‡ | 2.9 (-1.0, 6.9) | 5.3 (1.3, 9.5) |

**Legend:** Mean trajectory is centred on the sex-specific mean of the child (male mean: 0.0098 (SD:0.0008), and adult (male mean: 0.0076 (SD:0.0004) GRS. The difference in fat mass per SD higher child or adult GRS is back transformed from the log scale for ease of interpretation and is a ratio of geometric means, expressed as a percentage difference. \* Mean fat mass at age specified. † Percentage change per year in fat mass. ‡ Percentage difference in fat mass at age specified per SD higher child or adult GRS. § Percentage difference in change per year, per SD higher child or adult GRS. GRS, genetic risk score; SD, standard deviation; CI, confidence interval.

Compared with a female with mean childhood and adulthood GRS, a 1 SD higher childhood GRS or adulthood GRS was associated with 10.5% (95% CI: 8.6, 12.4) and 5.1% (95% CI: 3.2, 6.9) higher total fat mass at 9y, respectively (**Table 3** and **Figure 1B**). Results for the association of each GRS with rates of change in total fat mass in females were broadly similar to males, demonstrating no strong associations of either GRS with change in total fat mass across most spline periods, except for associations with change in total fat mass from 13 to 15y; a 1 SD higher childhood and adulthood GRS was associated with -1.1% (95% CI: -1.9, -0.3) and -0.9% (95% CI: -1.7, -0.2) per year slower rates of increase in total fat mass, respectively, from 13-15y. This resulted in a slight reduction in the childhood and adulthood GRS associations with total fat mass at 15 and 18y, though the childhood GRS associations remained stronger than the adulthood GRS associations at both ages; 7.5% (95% CI: 5.2, 9.8) and 4.4% (95% CI: 2.2, 6.7) higher total fat mass at 18y per SD higher childhood and adulthood GRS, respectively. At age 25y, the childhood and adulthood GRS were similarly associated with total fat mass in females, driven by slightly slower gains per year in total fat mass from 18 to 25y per SD higher childhood GRS.

**Table 3.**
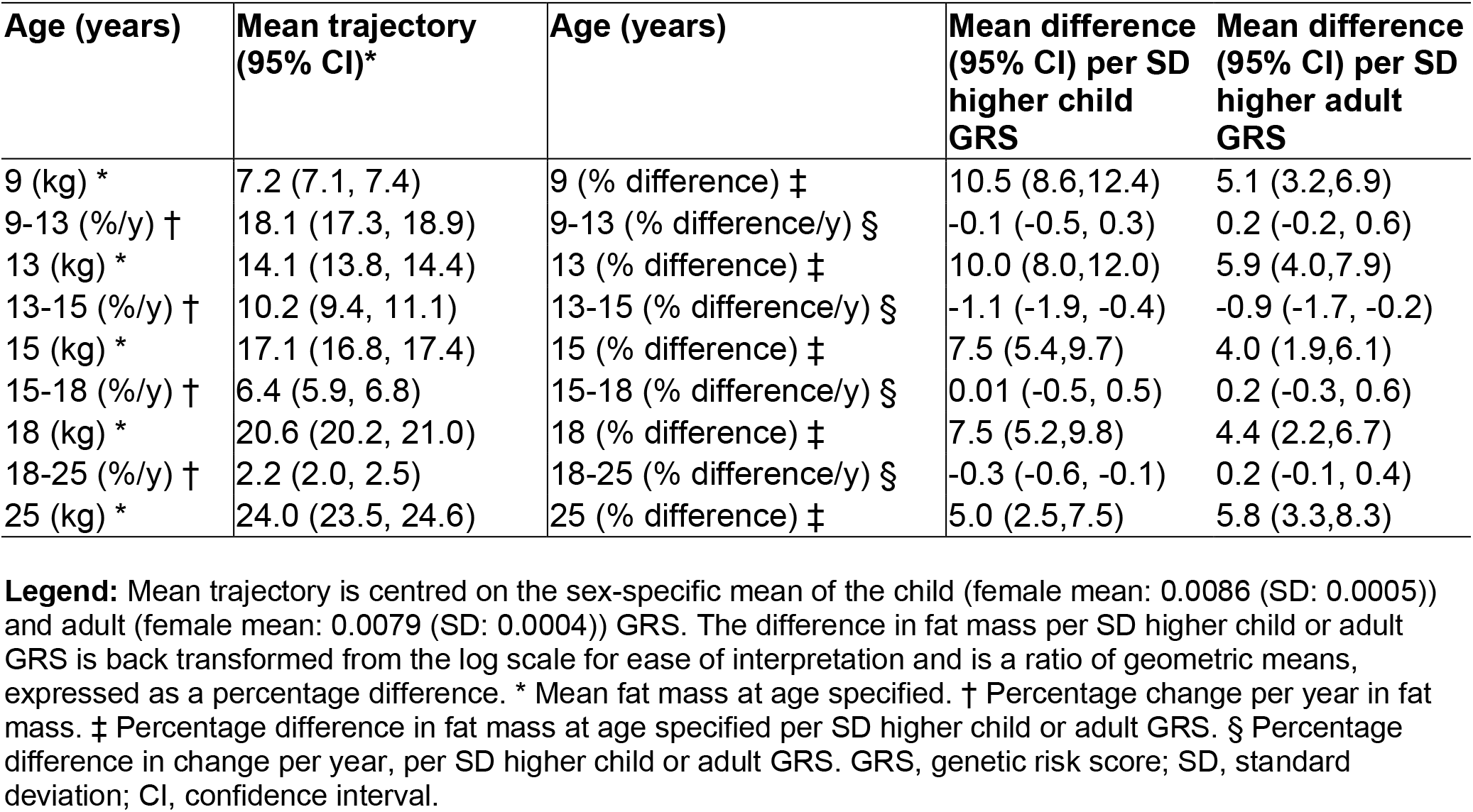
Mean trajectory and mean difference in trajectory of total fat mass from 9 to 25y per SD higher child GRS and adult GRS in females, with mutual adjustment for each score

| Age (years) | Mean trajectory (95% CI)* | Age (years) | Mean difference (95% CI) per SD higher child GRS | Mean difference (95% CI) per SD higher adult GRS |
| --- | --- | --- | --- | --- |
| 9 (kg) * | 7.2 (7.1, 7.4) | 9 (% difference) ‡ | 10.5 (8.6,12.4) | 5.1 (3.2,6.9) |
| 9-13 (%/y) † | 18.1 (17.3, 18.9) | 9-13 (% difference/y) § | -0.1 (-0.5, 0.3) | 0.2 (-0.2, 0.6) |
| 13 (kg) * | 14.1 (13.8, 14.4) | 13 (% difference) ‡ | 10.0 (8.0,12.0) | 5.9 (4.0,7.9) |
| 13-15 (%/y) † | 10.2 (9.4, 11.1) | 13-15 (% difference/y) § | -1.1 (-1.9, -0.4) | -0.9 (-1.7, -0.2) |
| 15 (kg) * | 17.1 (16.8, 17.4) | 15 (% difference) ‡ | 7.5 (5.4,9.7) | 4.0 (1.9,6.1) |
| 15-18 (%/y) † | 6.4 (5.9, 6.8) | 15-18 (% difference/y) § | 0.01 (-0.5, 0.5) | 0.2 (-0.3, 0.6) |
| 18 (kg) * | 20.6 (20.2, 21.0) | 18 (% difference) ‡ | 7.5 (5.2,9.8) | 4.4 (2.2,6.7) |
| 18-25 (%/y) † | 2.2 (2.0, 2.5) | 18-25 (% difference/y) § | -0.3 (-0.6, -0.1) | 0.2 (-0.1, 0.4) |
| 25 (kg) * | 24.0 (23.5, 24.6) | 25 (% difference) ‡ | 5.0 (2.5,7.5) | 5.8 (3.3,8.3) |

**Legend:** Mean trajectory is centred on the sex-specific mean of the child (female mean: 0.0086 (SD: 0.0005)) and adult (female mean: 0.0079 (SD: 0.0004)) GRS. The difference in fat mass per SD higher child or adult GRS is back transformed from the log scale for ease of interpretation and is a ratio of geometric means, expressed as a percentage difference. \* Mean fat mass at age specified. † Percentage change per year in fat mass. ‡ Percentage difference in fat mass at age specified per SD higher child or adult GRS. § Percentage difference in change per year, per SD higher child or adult GRS. GRS, genetic risk score; SD, standard deviation; CI, confidence interval.

Unadjusted findings (from models that did not adjust for the other GRS) were broadly similar to adjusted findings among males and females (**Supplementary Tables 2-3**). Mutually adjusted models using sex-combined GRS were also used. In males, a greater effect of the childhood GRS was observed in these models at earlier ages, unlike the sex specific GRS results. For example at age 9y a 1SD higher childhood GRS was associated with 14.9% (95% CI: 12.4, 17.5) higher fat mass (see **Supplementary Table 4**) compared to the 8.8% (95% CI 6.3, 11.3) difference seen in the sex specific model. In females, these models produced similar findings to our main models, albeit with slightly higher point estimates(**Supplementary Table 5**). Associations for childhood and adulthood GRS scores with trunk and peripheral fat mass were similar to our main findings on total fat mass (**Supplementary Figure 2**).

### Estimated effects of childhood and adulthood body size GRSs on trajectories of total lean mass

**Supplementary Figure 1.C-D** shows the mean sex-specific trajectories of total lean mass from 9 to 25y for 1) participants with the mean childhood GRS and mean adulthood GRS, 2) participants with a 1SD higher childhood GRS compared with the mean and 3) participants with a 1SD higher adulthood GRS compared with the mean. Compared with a male with mean childhood and adulthood GRS, a 1 SD higher childhood and adulthood GRS was associated with small differences in total lean mass at age 9y: 0.9% (95% CI: 0.4, 1.3) and 0.5% (95% CI: 0.03, 0.9) per SD higher childhood and adulthood GRS, respectively (**Supplementary Table 6** and **Figure 1.C**). The childhood GRS was associated with small differences in rates of change in total lean mass from 9 to 25y that differed in direction across spline periods whereas the adulthood GRS was associated with similar rates of change in total lean mass from 9-25y. By age 25y, a 1 SD higher childhood and adulthood GRS was associated with 1.1% (95% CI: 0.3, 2.0) and 1.4% (95% CI: 0.5, 2.3) higher total lean mass, respectively, among males. Findings were broadly similar for females (**Supplementary Table 7** and **Figure 1.D**). Unadjusted findings (from models that did not adjust for the other GRS score) and sex-combined GRS results were broadly similar to adjusted findings, see **Supplementary Tables 8-9** for unadjusted results and **Supplementary Tables 10-11** for mutually adjusted results using the sex-combined GRS.

## Discussion

In this prospective cohort study, we aimed to estimate the effects of life stage-specific genetic susceptibility to higher body size on trajectories of objectively measured body fat and lean mass from childhood to young adulthood. Our findings show that genetic variants for body size are more strongly associated with adiposity than lean mass from childhood to early adulthood. They also demonstrate that childhood variants are more strongly associated with adiposity in females than adulthood variants until early adulthood whereas childhood and adulthood variants are similarly associated with adiposity across early life in males. Our findings may inform selection of instruments for life stage-specific adiposity in future MR studies^25^.

Recently, a number of GWAS have been run in the Norwegian Mother, Father and Child Cohort Study (MoBa) between birth and 8 years of age, identifying novel SNPs which are associated with BMI specifically at different time points throughout this age range, suggesting that the genetic drivers of BMI change throughout early childhood^26^. Khera et al previously derived a genome-wide polygenic score (GPS) for BMI, using adult BMI-linked SNPs, combining a large set of SNPs with small effect sizes^27^. In that study, repeated measures of weight from ALSPAC, recorded from birth to 18 years of age, were used to establish that this GPS for adult BMI is associated with increased body weight already in early childhood, with associations further strengthening into young adulthood. However, little is known about whether distinct genetic influences exist on childhood vs. adulthood adiposity.

In our present study, we used a smaller set of SNPs which were strongly associated (P<5×10^−8^) with generally measured ‘body size’ to estimate their effects on more objective measures of body fat and lean mass at different stages of the early life course. This was important because the source GWAS for these SNPs, whilst substantially larger than other GWAS of childhood BMI, used a self-reported measure of recalled body size relative to peers when aged 10 years (measured via questionnaire several decades later). Despite these variants having been used in several MR studies to instrument ‘body size’ in childhood, it is unknown whether these variants are truly instrumenting adiposity as opposed to lean mass, how these variants effect trajectories of fat and lean mass throughout childhood, and whether these effects differ by sex. Our results suggest that genetic variants for body size are more strongly associated with body fat than lean mass across early life in both sexes. Moreover, our findings show that in females during childhood and adolescence, the childhood body size SNPs are more strongly associated with adiposity than the adulthood SNPs whereas childhood and adulthood SNPs are similarly associated with adiposity in males from childhood to early adulthood. Gaining a better understanding of the life stage- and sex-specific genomic components of adiposity may aid in the development of targeted intervention strategies in the future. Such measures may include the genotyping of children at an early age, for the monitoring of genetic risk throughout generations, and the stratification of health advice. Extending our study in the future to later adulthood would further improve understanding of whether genetic risk of childhood adiposity influences body composition in later life, and precisely when the genetic variants for adulthood body size establish themselves as more greatly influential for adiposity.

The limitations of this study include the restriction of analyses to participants of white ethnicity (given the European sample from which genetic variants were discovered) and therefore generalisability of our findings other ethnic groups^29^. The DXA measures used to quantify fat and lean mass are considered superior to highly indirect measures such as BMI or waist circumference, but DXA measures are less precise than MRI^30,31^ and are unable to measure ectopic fat and may not have uniform precision across the range of BMI values^32^. ALSPAC data are longitudinal and there are potential issues arising from loss to follow up over time which is unlikely to be at random^33^; this may bias results, and we note that individuals included in our analysis are from higher socioeconomic backgrounds than those not included. Our exposures of interest (genetic risk scores) are randomly allocated (biologically), however, and as such this is unlikely to be a concern.

### Conclusions

Genetic variants for body size are more strongly associated with adiposity than lean mass from childhood to early adulthood; childhood variants are more strongly associated with adiposity in females until early adulthood whereas childhood and adulthood variants are similarly associated with adiposity across early life in males. Findings may inform selection of instruments for life stage specific adiposity in future Mendelian randomization studies.

## Supporting information

Supplemental File 1 - STROBE

## Data Availability

Individual-level ALSPAC data are available following an application. This process of managed access is detailed at www.bristol.ac.uk/alspac/researchers/access. Cohort details and data descriptions for ALSPAC are publicly available at the same web address.

## Acknowledgements

We are extremely grateful to all the families who took part in this study, the midwives for their help in recruiting them, and the whole ALSPAC team, which includes interviewers, computer and laboratory technicians, clerical workers, research scientists, volunteers, managers, receptionists and nurses.

## Data availability

Individual-level ALSPAC data are available following an application. This process of managed access is detailed at www.bristol.ac.uk/alspac/researchers/access.

Cohort details and data descriptions for ALSPAC are publicly available at the same web address.

## Author contributions

SW, LMOK, and JAB planned the study. SW conducted analyses and wrote the first draft. TGR, GDS, LMOK and JAB critically reviewed the intellectual content of manuscript drafts and SW approved the final version for submission.

## References

1. Conolly, A. & Davies, B. Health survey for England 2017—adult and child overweight and obesity. NHS Digital, NHS: Leeds, UK (2018).

2. Lin, W.-Y., Wang, Y.-C., Teng, I.-H., Liu, C. & Lou, X.-Y. Associations of five obesity metrics with epigenetic age acceleration: Evidence from 2,474 Taiwan Biobank participants. Obesity 29, 1731–1738 (2021).

3. Kresovich, J. et al. Associations of Body Composition and Physical Activity Level With Multiple Measures of Epigenetic Age Acceleration, Am. J. Epidemiol, 190, 984–993 (2021).

4. Weihrauch-Blüher, S., Schwarz, P. & Klusmann, J.-H. Childhood obesity: increased risk for cardiometabolic disease and cancer in adulthood. Metabolism 92, 147–152 (2019).

5. Lassale, C. et al. Separate and combined associations of obesity and metabolic health with coronary heart disease: a pan-European case-cohort analysis. Eur. Heart J. 39, 397–406 (2018).

6. Lee, D. H. et al. Comparison of the association of predicted fat mass, body mass index, and other obesity indicators with type 2 diabetes risk: two large prospective studies in US men and women. Eur. J. Epidemiol. 33, 1113–1123 (2018).

7. Marchesini, G., Moscatiello, S., Di Domizio, S. & Forlani, G. Obesity-associated liver disease. J. Clin. Endocrinol. Metab. 93, S74–80 (2008).

8. Powell-Wiley, T. M. et al. Neighborhood-level socioeconomic deprivation predicts weight gain in a multi-ethnic population: longitudinal data from the Dallas Heart Study. Prev. Med. 66, 22–27 (2014).

9. Kopp, W. How Western Diet And Lifestyle Drive The Pandemic Of Obesity And Civilization Diseases. Diabetes Metab. Syndr. Obes. 12, 2221–2236 (2019).

10. Speakman, J. R., Loos, R. J. F., O’Rahilly, S., Hirschhorn, J. N. & Allison, D. B. GWAS for BMI: a treasure trove of fundamental insights into the genetic basis of obesity. Int. J. Obes. 42, 1524–1531 (2018).

11. Pulit, S. L. et al. Meta-analysis of genome-wide association studies for body fat distribution in 694 649 individuals of European ancestry. Hum. Mol. Genet. 28, 166–174 (2019).

12. Bradfield, J. P. et al. A trans-ancestral meta-analysis of genome-wide association studies reveals loci associated with childhood obesity. Hum. Mol. Genet. 28, 3327–3338 (2019).

13. Richardson, T. G., Sanderson, E., Elsworth, B., Tilling, K. & Davey Smith, G. Use of genetic variation to separate the effects of early and later life adiposity on disease risk: mendelian randomisation study. BMJ 369, m1203 (2020).

14. Fraser, A. et al. Cohort Profile: the Avon Longitudinal Study of Parents and Children: ALSPAC mothers cohort. Int. J. Epidemiol. 42, 97–110 (2013).

15. Boyd, A. et al. Cohort profile: the ‘children of the 90s’—the index offspring of the Avon Longitudinal Study of Parents and Children. Int. J. Epidemiol. 42, 111–127 (2013).

16. Northstone, K. et al. The Avon Longitudinal Study of Parents and Children (ALSPAC): an update on the enrolled sample of index children in 2019. Wellcome Open Res 4, 51 (2019).

17. Harris, P. A. et al. Research electronic data capture (REDCap)--a metadata-driven methodology and workflow process for providing translational research informatics support. J. Biomed. Inform. 42, 377–381 (2009).

18. Tilling, K., Macdonald-Wallis, C., Lawlor, D. A., Hughes, R. A. & Howe, L. D. Modelling childhood growth using fractional polynomials and linear splines. Ann. Nutr. Metab. 65, 129–138 (2014).

19. O’Keeffe, L. M. et al. Sex-specific trajectories of measures of cardiovascular health during childhood and adolescence: A prospective cohort study. Atherosclerosis 278, 190–196 (2018).

20. O’Keeffe, L. M. et al. Data on trajectories of measures of cardiovascular health in the Avon Longitudinal Study of Parents and Children (ALSPAC). Data Brief 23, 103687 (2019).

21. O’Keeffe, L. M., Fraser, A. & Howe, L. D. Accounting for height in indices of body composition during childhood and adolescence. Wellcome Open Res. 4, 105 (2019).

22. Charlton, C., Rasbash, J., Browne, W. J., Healy, M. & Cameron, B. MLwiN Version 3.00. Centre for multilevel modelling, University of Bristol (2017).

23. Zhang, Z., Parker, R. M. A., Charlton, C. M. J., Leckie, G. & Browne, W. J. R2MLwiN: A Package to Run MLwiN from within R. Journal of Statistical Software, Articles 72, 1–43 (2016).

24. Frysz, M., Howe, L. D., Tobias, J. H. & Paternoster, L. Using SITAR (SuperImposition by Translation and Rotation) to estimate age at peak height velocity in Avon Longitudinal Study of Parents and Children. Wellcome Open Res 3, 90 (2018).

25. Davies, N. M., Holmes, M. V. & Davey Smith, G. Reading Mendelian randomisation studies: a guide, glossary, and checklist for clinicians. BMJ 362, k601 (2018).

26. Helgeland, Ø. et al. Characterization of the genetic architecture of infant and early childhood body mass index. Nat Metab 4, 344–358 (2022).

27. Khera, A. V. et al. Polygenic Prediction of Weight and Obesity Trajectories from Birth to Adulthood. Cell 177, 587–596.e9 (2019).

28. Loomba-Albrecht, L. A. & Styne, D. M. Effect of puberty on body composition. Curr. Opin. Endocrinol. Diabetes Obes. 16, 10–15 (2009).

29. Roberts, M. C., Khoury, M. J. & Mensah, G. A. Perspective: The Clinical Use of Polygenic Risk Scores: Race, Ethnicity, and Health Disparities. Ethn. Dis. 29, 513–516 (2019).

30. Karlsson, A.-K. et al. Measurements of total and regional body composition in preschool children: A comparison of MRI, DXA, and anthropometric data. Obesity 21, 1018–1024 (2013).

31. Lee, V. et al. Estimation of visceral fat in 9-to 13-year-old girls using dual-energy X-ray absorptiometry (DXA) and anthropometry. Obes Sci Pract 4, 437–447 (2018).

32. Meredith-Jones, K., Haszard, J., Stanger, N. & Taylor, R. Precision of DXA-Derived Visceral Fat Measurements in a Large Sample of Adults of Varying Body Size. Obesity 26, 505–512 (2018).

33. Wolke, D. et al. Selective drop-out in longitudinal studies and non-biased prediction of behaviour disorders. Br. J. Psychiatry 195, 249–256 (2009).

