## Supplementary material for "Life stage-specific effects of genetic susceptibility to higher body size on body fat and lean mass: prospective cohort study"

**SUPPLEMENTARY ANALYSES**

| **Supplementary Table 1** – Characteristics of participants not included in study due to missing genetic and/or adiposity data | | |
| --- | --- | --- |
|  | **Females**  **N = 2479** | **Males**  **N = 2805** |
|  | **n (%)** | **n (%)** |
| **Household social class*** |  |  |
| Professional | 97 (8.6) | 146 (10.4) |
| Managerial & Technical | 393 (35.0) | 529 (37.7) |
| Non-Manual | 297 (26.5) | 349 (24.9) |
| Manual | 231 (20.6) | 263 (18.8) |
| Part skilled & unskilled | 105 (9.4) | 116 (8.3) |
| **Mother’s highest educational qualification†** |  |  |
| Less than O level | 543 (42.4) | 691 (43.0) |
| O level | 440 (34.4) | 524 (32.6) |
| A level | 202 (15.8) | 254 (15.8) |
| Degree or above | 96 (7.5) | 137 (8.5) |
| **Mother’s partner’s highest educational qualification†** |  |  |
| Less than O level | 569 (46.9) | 691 (45.8) |
| O level | 236 (19.5) | 293 (19.4) |
| A level | 282 (23.3) | 329 (21.8) |
| Degree or above | 126 (10.4) | 197 (13.1) |
| **Maternal smoking during pregnancy (first three months)‡** | 524 (34.4) | 657 (34.7) |
|  | **Mean (SD)** | **Mean (SD)** |
| **Birth weight (kg)** | 3.25 (0.61) | 3.34 (0.67) |
| **Legend:** The total N for these characteristics differs somewhat from sample sizes used in analyses as these complete data on these characteristics was not required for our analyses..  DXA - dual-energy X-ray absorptiometry.  *****Household social class was measured as the highest of the mother’s or her partner’s occupational social class using data on job title and details of occupation collected about the mother and her partner from the mother’s questionnaire at 32 weeks gestation. Social class was derived using the standard occupational classification (SOC) codes developed by the United Kingdom Office of Population Census and Surveys and classified as I-professional, II-managerial and technical, IIINM-non-manual, IIIM-manual, and IV&V part skilled occupations and unskilled occupations.  **†**Education status was recorded via a questionnaire of the mother, asking for her highest level of education, and her partner's highest level of education at 32 weeks of gestation. Responses are categorised into less than O-level (including vocational courses and the certificate of secondary education (CSE), O-level (taken at 16 years), A-levels (taken at 18 years), or university degree (or higher).  **‡**Maternal smoking in the first trimester was self-reported via questionnaire of the mothers; form of smoking was defined as: cigarettes, cigars, pipe, or ‘other’, all of which we combine into a binarized value of having smoked or not having smoked during pregnancy. | | |

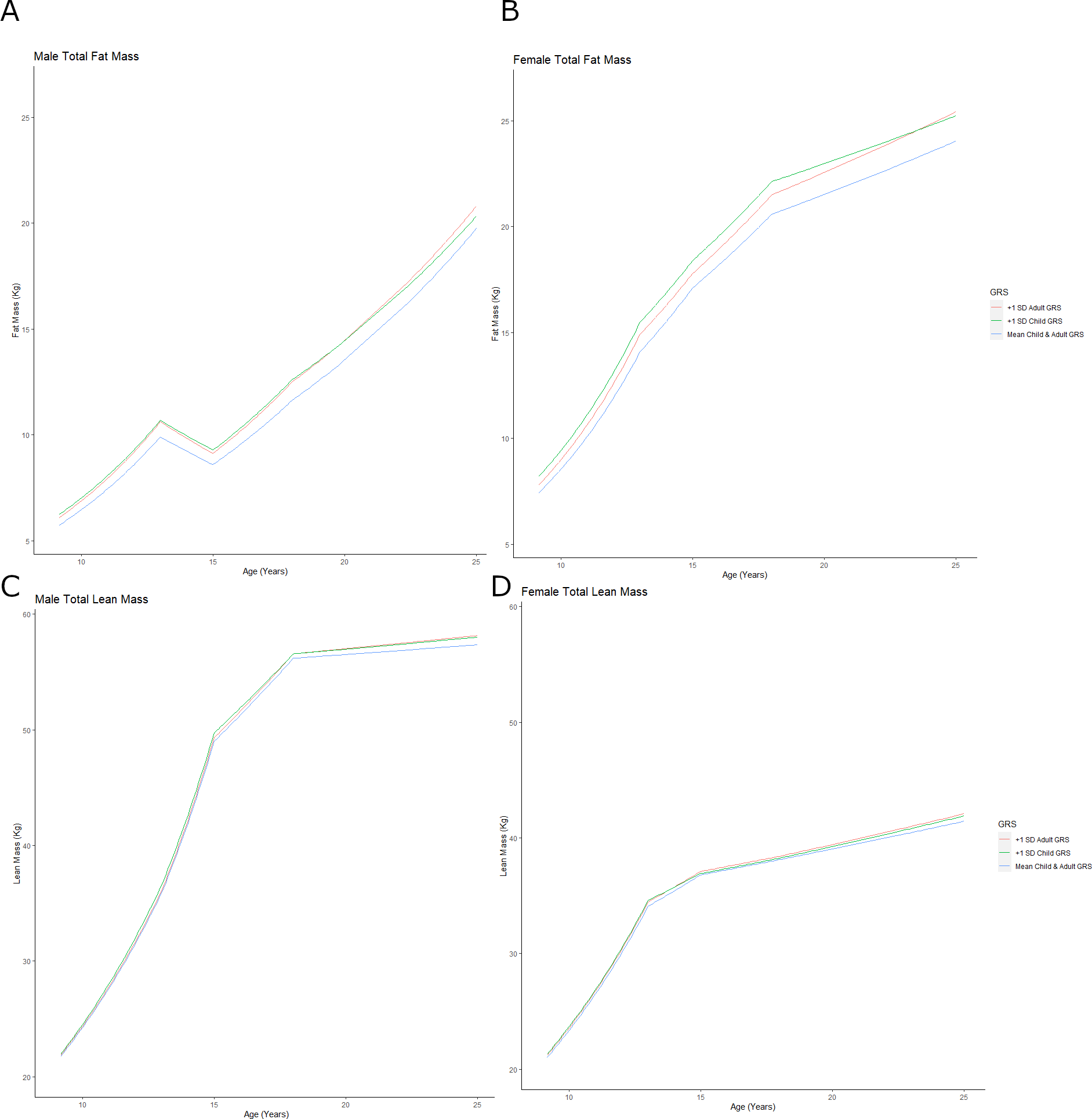

**Supplementary Figure 1** - Mean trajectories of fat (A, B) and lean mass (C, D) in females and males from 9 to 25 years by child and adult GRS scores. Trajectories are shown for mean childhood and adulthood GRS, along with trajectories for a participant with a 1 SD higher childhood GRS than the mean and 1 SD higher adulthood GRS than the mean. Trajectories derived from models which include mutual adjustment for each score

| **Supplementary Table 2** - Mean trajectory and mean difference in trajectory of total fat mass from 9 to 25y per SD higher child GRS and adult GRS in males, from models without mutual adjustment for the other score | | | | |
| --- | --- | --- | --- | --- |
| **Age (years)** | **Mean trajectory (95% CI)*** | **Age (years)** | **Mean difference (95% CI) per SD higher child GRS** | **Mean difference (95% CI) per SD higher adult GRS** |
| 9 (kg) * | 5.6 (5.4, 5.8) | 9 (% difference) ‡ | 11.3 (9.0, 13.7) | 9.6 (7.3, 12.0) |
| 9-13 (%/y) † | 13.3 (13.1, 13.6) | 9-13 (% difference/y) § | 0.3 (0.2, 0.5) | 0.3 (0.1, 0.4) |
| 13 (kg) * | 9.9 (9.7, 10.2) | 13 (% difference) ‡ | 10.8 (8.3, 13.4) | 10.4 (7.9,13.0) |
| 13-15 (%/y) † | 19.1 (18.8, 19.4) | 13-15 (% difference/y) § | -0.03 (-0.25, 0.20) | 0.2 (-0.1, 0.4) |
| 15 (kg) * | 8.6 (8.4, 8.8) | 15 (% difference) ‡ | 10.5 (7.8, 13.2) | 9.3 (6.6, 12.0) |
| 15-18 (%/y) † | 6.4 (6.2, 6.6) | 15-18 (% difference/y) § | -0.3 (-0.5, -0.1) | 0.01 (-0.2, 0.2) |
| 18 (kg) * | 11.6 (11.3, 12.0) | 18 (% difference) ‡ | 11.4 (8.3, 14.5) | 10.9 (7.8, 14.0) |
| 18-25 (%/y) † | -0.03, (-0.1, 0.04) | 18-25 (% difference/y) § | -0.1 (-0.2, -0.02) | -0.2 (-0.2, -0.1) |
| 25 (kg) * | 19.8 (19.1, 20.5) | 25 (% difference) ‡ | 4.9 (1.3, 8.7) | 6.3 (2.6, 10.1) |
| **Legend:** Mean trajectory is centred on the sex-specific mean of the child (male mean: 0.0098 (SD:0.0008), and adult (male mean: 0.0076 (SD:0.0004) GRS.. The difference in fat mass per SD higher child or adult GRS is back transformed from the log scale for ease of interpretation and is a ratio of geometric means, expressed as a percentage difference. * Mean fat mass at age specified. † Percentage change per year in fat mass. ‡ Percentage difference in fat mass at age specified per SD higher child or adult GRS. § Percentage difference in change per year, per SD higher child or adult GRS. GRS, genetic risk score; SD, standard deviation; CI, confidence interval. | | | | |

| **Supplementary Table 3** - Mean trajectory and mean difference in trajectory of total fat mass from 9 to 25y per SD higher child GRS and adult GRS in females, from models without mutual adjustment for the other score | | | | |
| --- | --- | --- | --- | --- |
| **Age (years)** | **Mean trajectory (95% CI)*** | **Age (years)** | **Mean difference (95% CI) per SD higher child GRS** | **Mean difference (95% CI) per SD higher adult GRS** |
| 9 (kg) * | 7.2 (7.1, 7.4) | 9 (% difference) ‡ | 12.5 (10.7, 14.4) | 9.0 (7.2, 10.9) |
| 9-13 (%/y) † | 18..0 (17.7, 18.3) | 9-13 (% difference/y) § | 0.3 (0.2, 0.4) | 0.3 (0.1, 0.4) |
| 13 (kg) * | 14.1 (13.8, 14.4) | 13 (% difference) ‡ | 12.4 (10.5, 14.3) | 9.7 (7.9, 11.7) |
| 13-15 (%/y) † | 10.7 (10.4, 11.0) | 13-15 (% difference/y) § | -0.5 (-0.8, 0.3) | -0.4 (-0.6, -0.1) |
| 15 (kg) * | 17.1 (16.8, 17.4) | 15 (% difference) ‡ | 9.1 (7.1, 11.1) | 6.8 (4.8, 8.8) |
| 15-18 (%/y) † | 2.4 (2.2, 2.5) | 15-18 (% difference/y) § | -0.04 (0.2, 0.1) | 0.1 (-0.1, 0.3) |
| 18 (kg) * | 20.6 (20.2, 21.0) | 18 (% difference) ‡ | 9.2 (7.1, 11.4) | 7.3 (5.1, 9.5) |
| 18-25 (%/y) † | -1.0 (-1.0, -0.9) | 18-25 (% difference/y) § | -0.2 (-0.3, -0.2) | -0.1 (-0.2, -0.04) |
| 25 (kg) * | 24.0 (23.5, 24.6) | 25 (% difference) ‡ | 7.2 (4.8, 9.6) | 7.8 (5.4, 10.2) |
| **Legend:** Mean trajectory is centred on the sex-specific mean of the child (female mean: 0.0086 (SD: 0.0005)) and adult (female mean: 0.0079 (SD: 0.0004)) GRS. The difference in fat mass per SD higher child or adult GRS is back transformed from the log scale for ease of interpretation and is a ratio of geometric means, expressed as a percentage difference. * Mean fat mass at age specified. † Percentage change per year in fat mass. ‡ Percentage difference in fat mass at age specified per SD higher child or adult GRS. § Percentage difference in change per year, per SD higher child or adult GRS. GRS, genetic risk score | | | | |

| **Supplementary Table 4 -** Mean trajectory and mean difference in trajectory of total fat mass from 9 to 25y per SD higher child GRS and adult GRS in (using sex-combined GRS scores) in males, with mutual adjustment for each score | | | | |
| --- | --- | --- | --- | --- |
| **Age (years)** | **Mean trajectory (95% CI)*** | **Age (years)** | **Mean difference (95% CI) per SD higher child GRS** | **Mean difference (95% CI)**  **per SD higher adult GRS** |
| 9 (kg) * | 5.6 (5.4, 5.8) | 9 (% difference) ‡ | 14.9 (12.4, 17.5) | 4.8 (2.5, 7.2) |
| 9-13 (%/y) † | 15.3 (14.2, 16.5) | 9-13 (% difference/y) § | -0.3 (-0.8, 0.3) | 0.3 (-0.3, 0.8) |
| 13 (kg) * | 9.9 (9.7,10.2) | 13 (% difference) ‡ | 13.6 (11.0, 16.4) | 5.9 (3.4, 8.5) |
| 13-15 (%/y) † | -6.9 (-7.9, -5.9) | 13-15 (% difference/y) § | 0.1 (-0.9, 1.1) | -0.1 (-1.1, 0.9) |
| 15 (kg) * | 8.6 (9.7, 10.2) | 15 (% difference) ‡ | 13.8 (11.0, 16.7) | 5.6 (3.0, 8.4) |
| 15-18 (%/y) † | 10.6 (9.8, 11.4) | 15-18 (% difference/y) § | -0.4 (-1.2, 0.3) | 1.1 (0.4, 1.9) |
| 18 (kg) * | 11.6 (11.3, 12.0) | 18 (% difference) ‡ | 12.3 (9.1, 15.7) | 9.3 (6.2, 12.6) |
| 18-25 (%/y) † | 7.8 (7.4, 8.3) | 18-25 (% difference/y) § | -1.0 (-1.4, -0.6) | -0.2 (-0.6, 0.2) |
| 25 (kg) * | 19.7 (19.0, 20.4) | 25 (% difference) ‡ | 4.6 (0.8, 8.6) | 7.6 (3.8, 11.6) |
| **Legend:** Mean trajectory is centred on the sex-specific mean of the child (male mean: 0.0098 (SD:0.0008), and adult (male mean: 0.0076 (SD:0.0004) GRS. The difference in fat mass per SD higher child or adult GRS is back transformed from the log scale for ease of interpretation and is a ratio of geometric means, expressed as a percentage difference. * Mean fat mass at age specified. † Percentage change per year in fat mass. ‡ Percentage difference in fat mass at age specified per SD higher child or adult GRS. § Percentage difference in change per year, per SD higher child or adult GRS. GRS, genetic risk score; SD, standard deviation; CI, confidence interval. | | | | |

| **Supplementary Table 5 -** Mean trajectory and mean difference in trajectory of total fat mass from 9 to 25y per SD higher child GRS and adult GRS in (using sex-combined GRS scores) in females, with mutual adjustment for each score | | | | |
| --- | --- | --- | --- | --- |
| **Age (years)** | **Mean trajectory (95% CI)*** | **Age (years)** | **Mean difference (95% CI) per SD higher child GRS** | **Mean difference (95% CI) per SD higher adult GRS** |
| 9 (kg) * | 7.2 (7.1, 7.4) | 9 (% difference) ‡ | 12.9 (10.9, 14.9) | 6.3 (4.5, 8.2) |
| 9-13 (%/y) † | 18.1 (17.3, 19.0) | 9-13 (% difference/y) § | -0.2 (-0.6, 0.2) | 0.3 (-0.1, 0.6) |
| 13 (kg) * | 14.1 (13.8, 14.4) | 13 (% difference) ‡ | 11.9 (9.94, 14.0) | 7.4 (5.5, 9.4) |
| 13-15 (%/y) † | 10.2 (9.4, 11.1) | 13-15 (% difference/y) § | -1.4 (-2.1, -0.6) | -0.9 (-1.6, -0.2) |
| 15 (kg) * | 17.1 (16.8, 17.4) | 15 (% difference) ‡ | 8.9 (6.8, 11.1) | 5.5 (3.5, 7.6) |
| 15-18 (%/y) † | 6.4 (5.9, 6.8) | 15-18 (% difference/y) § | -0.3 (-0.7, 0.2) | 0.5 (0.1, 1.0) |
| 18 (kg) * | 20.6 (20.2, 21.0) | 18 (% difference) ‡ | 8.1 (5.9, 10.4) | 7.2 (5.0, 9.4) |
| 18-25 (%/y) † | 2.3 (2.0, 2.5) | 18-25 (% difference/y) § | -0.3 (-0.6, -0.1) | 0.1 (-0.1, 0.3) |
| 25 (kg) * | 24.1 (23.5, 24.6) | 25 (% difference) ‡ | 5.5 (3.1, 8.0) | 7.9 (5.4, 10.4) |
| **Legend:** Mean trajectory is centred on the sex-specific mean of the child (female mean: 0.0086 (SD: 0.0005)) and adult (female mean: 0.0079 (SD: 0.0004)) GRS. . The difference in fat mass per SD higher child or adult GRS is back transformed from the log scale for ease of interpretation and is a ratio of geometric means, expressed as a percentage difference. * Mean fat mass at age specified. † Percentage change per year in fat mass. ‡ Percentage difference in fat mass at age specified per SD higher child or adult GRS. § Percentage difference in change per year, per SD higher child or adult GRS. GRS, genetic risk score; SD, standard deviation; CI, confidence interval. | | | | |

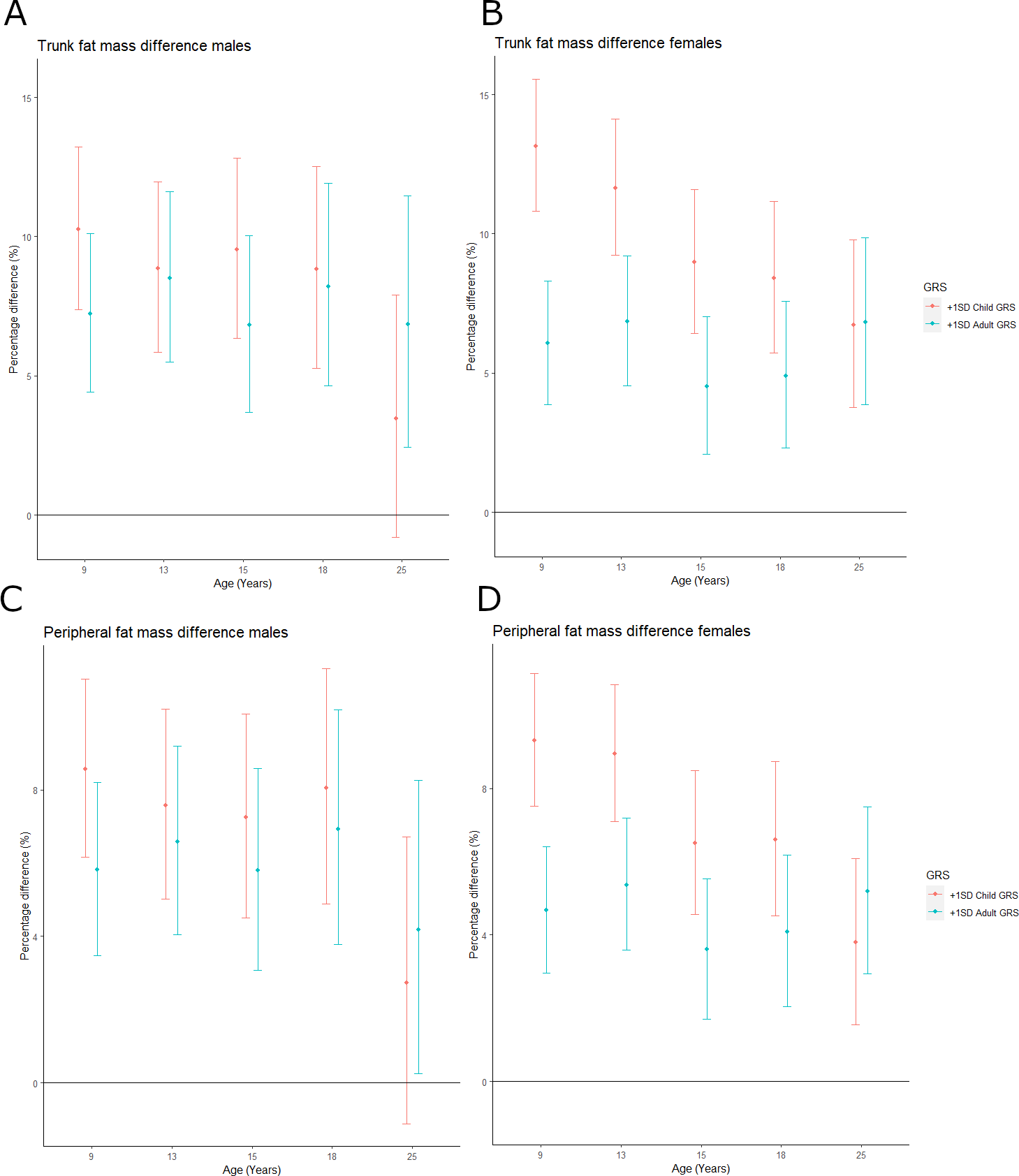

**Supplementary Figure 2 -** Trunk and peripheral fat mass differences**:** A/B - percentage difference between the mean childhood and adulthood GRS model and the +1SD GRS models for males (A) and females (B) of trunk fat mass from age 9 to 25. C/D - percentage difference between the mean childhood and adulthood GRS model and the +1SD GRS models for males (C) and females (D) of peripheral fat mass from age 9 to 25.

| **Supplementary Table 6** - Mean trajectory and mean difference in trajectory of total lean mass from 9 to 25y per SD higher child GRS and adult GRS in males, with mutual adjustment for each score | | | | |
| --- | --- | --- | --- | --- |
| **Age (years)** | **Mean trajectory (95% CI)*** | **Age (years)** | **Mean difference (95% CI) per SD higher child GRS** | **Mean difference (95% CI) per SD higher adult GRS** |
| 9 (kg) * | 21.3 (21.1, 21.4) | 9 (% difference) ‡ | 0.9 (0.4, 1.3) | 0.5 (0.03, 0.9) |
| 9-13 (%/y) † | 13.9 (13.6, 14.3) | 9-13 (% difference/y) § | 0.3 (0.1, 0.4) | 0.01 (-0.2, 0.2) |
| 13 (kg) * | 35.9 (35.6, 36.1) | 13 (% difference) ‡ | 1.9 (1.3, 2.5) | 0.5 (-0.1, 1.1) |
| 13-15 (%/y) † | 16.9 (16.6, 17.2) | 13-15 (% difference/y) § | -0.2 (-0.4, 0.1) | 0.1 (-0.2 ,0.3) |
| 15 (kg) * | 49.0 (48.7, 49.3) | 15 (% difference) ‡ | 1.5 (0.9, 2.1) | 0.7 (0.03, 1.3) |
| 15-18 (%/y) † | 4.7 (4.4, 4.9) | 15-18 (% difference/y) § | -0.3 (-0.5, -0.04) | 0.01 (-0.2, 0.2) |
| 18 (kg) * | 56.2 (55.8, 56.6) | 18 (% difference) ‡ | 0.7 (-0.1, 1.5) | 0.7 (-0.1, 1.5) |
| 18-25 (%/y) † | 0.3 (0.2, 0.4) | 18-25 (% difference/y) § | 0.1 (-0.02, 0.2) | 0.1 (0.01, 0.2) |
| 25 (kg) * | 57.4 (56.9, 57.8) | 25 (% difference) ‡ | 1.1 (0.3, 2.0) | 1.4 (0.5, 2.3) |
| **Legend:** Mean trajectory is centred on the sex-specific mean of the child (male mean: 0.0098 (SD:0.0008), and adult (male mean: 0.0076 (SD:0.0004) GRS. The difference in fat mass per SD higher child or adult GRS is back transformed from the log scale for ease of interpretation and is a ratio of geometric means, expressed as a percentage difference. * Mean fat mass at age specified. † Percentage change per year in fat mass. ‡ Percentage difference in fat mass at age specified per SD higher child or adult GRS. § Percentage difference in change per year, per SD higher child or adult GRS. GRS, genetic risk score; SD, standard deviation; CI, confidence interval. | | | | |

| **Supplementary Table 7** - Mean trajectory and mean difference in trajectory of total lean mass from 9 to 25y per SD higher child GRS and adult GRS in females, with mutual adjustment for each score | | | | |
| --- | --- | --- | --- | --- |
| **Age (years)** | **Mean trajectory (95% CI)*** | **Age (years)** | **Mean difference (95% CI) per SD higher child GRS** | **Mean difference (95% CI) per SD higher adult GRS** |
| 9 (kg) * | 20.6 (20.4, 20.7) | 9 (% difference) ‡ | 1.4 (0.9, 1.9) | 0.9 (0.4, 1.4) |
| 9-13 (%/y) † | 13.4 (13.2, 13.7) | 9-13 (% difference/y) § | 0.02 (-0.1, 0.1) | 0.04 (-0.1, 0.2) |
| 13 (kg) * | 34.1 (33.9, 34.3) | 13 (% difference) ‡ | 1.5 (1.0, 2.0) | 1.1 (0.6, 1.6) |
| 13-15 (%/y) † | 3.9 (3.7, 4.2) | 13-15 (% difference/y) § | -0.6 (-0.8, -0.3) | -0.2 (-0.4, 0.1) |
| 15 (kg) * | 36.8 (36.6, 37.0) | 15 (% difference) ‡ | 0.3 (-0.3, 0.9) | 0.8 (0.2, 1.4) |
| 15-18 (%/y) † | 1.2 (1.1, 1.3) | 15-18 (% difference/y) § | 0.01 (-0.1, 0.1) | -0.01 (-0.1, 0.1) |
| 18 (kg) * | 38.1 (37.9, 38.3) | 18 (% difference) ‡ | 0.3 (-0.2, 0.9) | 0.8 (0.2, 1.3) |
| 18-25 (%/y) † | 1.2 (1.1, 1.3) | 18-25 (% difference/y) § | 0.1 (0.1, 0.2) | 0.1 (0.04, 0.2) |
| 25 (kg) * | 41.4 (41.2, 41.7) | 25 (% difference) ‡ | 1.1 (0.5, 1.8) | 1.6 (0.9, 2.2) |
| **Legend:** Mean trajectory is centred on the sex-specific mean of the child (female mean: 0.0086 (SD: 0.0005)) and adult (female mean: 0.0079 (SD: 0.0004)) GRS.. The difference in fat mass per SD higher child or adult GRS is back transformed from the log scale for ease of interpretation and is a ratio of geometric means, expressed as a percentage difference. * Mean fat mass at age specified. † Percentage change per year in fat mass. ‡ Percentage difference in fat mass at age specified per SD higher child or adult GRS. § Percentage difference in change per year, per SD higher child or adult GRS. GRS, genetic risk score; SD, standard deviation; CI, confidence interval. | | | | |

| **Supplementary Table 8** - Mean trajectory and mean difference in trajectory of total lean mass from 9 to 25y per SD higher child GRS and adult GRS in males, without mutual adjustment for each score | | | | |
| --- | --- | --- | --- | --- |
| **Age (years)** | **Mean trajectory (95% CI)*** | **Age (years)** | **Mean difference (95% CI) per SD higher child GRS** | **Mean difference (95% CI) per SD higher adult GRS** |
| 9 (kg) * | 21.3 (21.1, 21.4) | 9 (% difference) ‡ | 1.0 (0.6, 1.5) | 0.8 (0.4, 1.2) |
| 9-13 (%/y) † | 13.3 (13.1, 13.6) | 9-13 (% difference/y) § | 0.3 (0.2, 0.5) | 0.3 (0.1, 0.4) |
| 13 (kg) * | 35.9 (35.6 36.1) | 13 (% difference) ‡ | 2.1 (1.5, 2.6) | 1.2 (0.7, 1.8) |
| 13-15 (%/y) † | 19.1 (18.8, 19.4) | 13-15 (% difference/y) § | -0.03, -0.3, 0.2) | 0.2, (0-.1, 0.4) |
| 15 (kg) * | 49.0 (48.7, 49.3) | 15 (% difference) ‡ | 1.7 (1.2, 2.3) | 1.2 (0.6, 1.8) |
| 15-18 (%/y) † | 6.4 (6.2, 6.6) | 15-18 (% difference/y) § | -0.3 (-0.5, -0.1) | 0.01 (-0.2, 0.2) |
| 18 (kg) * | 56.2 (55.8, 56.6) | 18 (% difference) ‡ | 0.9 (0.2, 1.7) | 0.9 (0.2, 1.7) |
| 18-25 (%/y) † | -0.03 (-0.1, 0.04) | 18-25 (% difference/y) § | -0.1 (-0.2, -0.02) | -0.2 (-0.2, -0.1) |
| 25 (kg) * | 57.3 (56.9, 57.8) | 25 (% difference) ‡ | 1.7 (0.9, 2.5) | 1.8 (1.0, 2.6) |
| **Legend:** Mean trajectory is centred on the sex-specific mean of the child (male mean: 0.0098 (SD:0.0008), and adult (male mean: 0.0076 (SD:0.0004) GRS. The difference in fat mass per SD higher child or adult GRS is back transformed from the log scale for ease of interpretation and is a ratio of geometric means, expressed as a percentage difference. * Mean fat mass at age specified. † Percentage change per year in fat mass. ‡ Percentage difference in fat mass at age specified per SD higher child or adult GRS. § Percentage difference in change per year, per SD higher child or adult GRS. GRS, genetic risk score; SD, standard deviation; CI, confidence interval. | | | | |

| **Supplementary Table 9** - Mean trajectory and mean difference in trajectory of total lean mass from 9 to 25y per SD higher child GRS and adult GRS in females, without mutual adjustment for each score | | | | |
| --- | --- | --- | --- | --- |
| **Age (years)** | **Mean trajectory (95% CI)*** | **Age (years)** | **Mean difference (95% CI) per SD higher child GRS** | **Mean difference (95% CI) per SD higher adult GRS** |
| 9 (kg) * | 20.6 (20.4, 20.7) | 9 (% difference) ‡ | 1.7 (1.3, 2.2) | 1.4 (1.0, 1.9) |
| 9-13 (%/y) † | 18.0 (17.7, 18.3) | 9-13 (% difference/y) § | 0.3 (0.2, 0.4) | 0.3 (0.1, 0.4) |
| 13 (kg) * | 34.1 (33.9, 34.3) | 13 (% difference) ‡ | 1.9 (1.4, 2.4) | 1.6 (1.2, 2.1) |
| 13-15 (%/y) † | 10.7 (10.4, 11.0) | 13-15 (% difference/y) § | -0.5 (-0.8, -0.3) | -0.4 (-0.6, -0.1) |
| 15 (kg) * | 36.8 (36.6, 37.0) | 15 (% difference) ‡ | 0.6 (0.1, 1.1) | 0.9 (0.4, 1.4) |
| 15-18 (%/y) † | 2.4 (2.2, 2.5) | 15-18 (% difference/y) § | -0.04 (-0.2, 0.1) | 0.1 (-0.1, 0.2) |
| 18 (kg) * | 38.1 (37.9, 38.3) | 18 (% difference) ‡ | 0.6 (0.1, 1.1) | 0.9 (0.4, 1.4) |
| 18-25 (%/y) † | -1.0 (-1.0, -0.9) | 18-25 (% difference/y) § | -0.2 (-0.3, -0.2) | -0.1 (-0.2, -0.04) |
| 25 (kg) * | 41.4 (41.2, 41.7) | 25 (% difference) ‡ | 1.7 (1.1, 2.3) | 2.0 (1.4, 2.6) |
| **Legend:** Mean trajectory is centred on the sex-specific mean of the child (female mean: 0.0086 (SD: 0.0005)) and adult (female mean: 0.0079 (SD: 0.0004)) GRS. The difference in fat mass per SD higher child or adult GRS is back transformed from the log scale for ease of interpretation and is a ratio of geometric means, expressed as a percentage difference. * Mean fat mass at age specified. † Percentage change per year in fat mass. ‡ Percentage difference in fat mass at age specified per SD higher child or adult GRS. § Percentage difference in change per year, per SD higher child or adult GRS. GRS, genetic risk score; SD, standard deviation; CI, confidence interval. | | | | |

| **Supplementary Table 10** - Mean trajectory and mean difference in trajectory of total lean mass from 9 to 25y per SD higher child GRS and adult GRS (using sex-combined GRS scores) in males, without mutual adjustment for each score | | | | |
| --- | --- | --- | --- | --- |
| **Age (years)** | **Mean trajectory (95% CI)*** | **Age (years)** | **Mean difference (95% CI) per SD higher child GRS** | **Mean difference (95% CI) per SD higher adult GRS** |
| 9 (kg) * | 21.3 (21.1, 21.4) | 9 (% difference) ‡ | 1.3 (0.8, 1.7) | 0.3 (-0.2, 0.7) |
| 9-13 (%/y) † | 13.9 (13.6, 14.3) | 9-13 (% difference/y) § | -0.3 (0.1, 0.4) | 0.2 (0.1, 0.4) |
| 13 (kg) * | 35.9 (35.6 36.1) | 13 (% difference) ‡ | 2.3 (1.7, 2.9) | 1.3 (0.7, 1.9) |
| 13-15 (%/y) † | 16.9 (16.6, 17.2) | 13-15 (% difference/y) § | -0.2 (-0.5, -0.01) | -0.1 (-0.3, 0.1) |
| 15 (kg) * | 49.0 (48.7, 49.3) | 15 (% difference) ‡ | 1.8 (1.2, 2.4) | 1.1 (0.5, 1.7) |
| 15-18 (%/y) † | 4.7 (4.4, 4.9) | 15-18 (% difference/y) § | -0.3 (-0,5, -0.1) | -0.1 (-0.3, 0.1) |
| 18 (kg) * | 56.2 (55.8, 56.6) | 18 (% difference) ‡ | 1.0 (0.2, 1.7) | 0.8 (0.1, 1.6) |
| 18-25 (%/y) † | 0.3 (0.2, 0.4) | 18-25 (% difference/y) § | 0.1 (0.05, 0.2) | 0.1 (-0.01, 0.2) |
| 25 (kg) * | 57.4 (56.9, 57.8) | 25 (% difference) ‡ | 2.0 (1.1, 2.8) | 1.3 (0.5, 2.2) |
| **Legend:** Mean trajectory is centred on the sex-specific mean of the child (male mean: 0.0098 (SD:0.0008), and adult (male mean: 0.0076 (SD:0.0004) GRS. The difference in fat mass per SD higher child or adult GRS is back transformed from the log scale for ease of interpretation and is a ratio of geometric means, expressed as a percentage difference. * Mean fat mass at age specified. † Percentage change per year in fat mass. ‡ Percentage difference in fat mass at age specified per SD higher child or adult GRS. § Percentage difference in change per year, per SD higher child or adult GRS. GRS, genetic risk score; SD, standard deviation; CI, confidence interval. | | | | |

| **Supplementary Table 11** - Mean trajectory and mean difference in trajectory of total lean mass from 9 to 25y per SD higher child GRS and adult GRS (using sex-combined GRS scores) in females, without mutual adjustment for each score | | | | |
| --- | --- | --- | --- | --- |
| **Age (years)** | **Mean trajectory (95% CI)*** | **Age (years)** | **Mean difference (95% CI) per SD higher child GRS** | **Mean difference (95% CI) per SD higher adult GRS** |
| 9 (kg) * | 20.6 (20.4, 20.7) | 9 (% difference) ‡ | 1.9 (1.4, 2.4) | 0.9 (0.4, 1.4) |
| 9-13 (%/y) † | 13.5 (13.2, 13.7) | 9-13 (% difference/y) § | 0.02 (-0.1, 0.1) | 0.1 (0.02, 0.2) |
| 13 (kg) * | 34.1 (33.9, 34.3) | 13 (% difference) ‡ | 2.0 (1.5, 2.5) | 1.4 (0.9, 1.9) |
| 13-15 (%/y) † | 3.9 (3.7, 4.2) | 13-15 (% difference/y) § | -0.6 (-0.8, -0.4) | -0.5 (-0.7, -0.2) |
| 15 (kg) * | 36.8 (36.6, 37.0) | 15 (% difference) ‡ | 0.8 (0.2, 1.3) | 0.5 (-0.1, 1.0) |
| 15-18 (%/y) † | 1.2 (1.1, 1.3) | 15-18 (% difference/y) § | -0.02 (0.1, 0.1) | 0.1 (-0.1, 0.2) |
| 18 (kg) * | 38.1 (37.9, 38.3) | 18 (% difference) ‡ | 0.7 (0.2, 1.3) | 0.7 (0.1, 1.2) |
| 18-25 (%/y) † | 1.3 (1.1, 1.3) | 18-25 (% difference/y) § | 0.1 (0.07, 0.2) | 0.1 (0.03, 0.2) |
| 25 (kg) * | 41.4 (41.2, 41.7) | 25 (% difference) ‡ | 1.7 (1.1, 2.3) | 1.4 (0.7, 2.0) |
| **Legend:** Mean trajectory is centred on the sex-specific mean of the child (female mean: 0.0086 (SD: 0.0005)) and adult (female mean: 0.0079 (SD: 0.0004)) GRS. The difference in fat mass per SD higher child or adult GRS is back transformed from the log scale for ease of interpretation and is a ratio of geometric means, expressed as a percentage difference. * Mean fat mass at age specified. † Percentage change per year in fat mass. ‡ Percentage difference in fat mass at age specified per SD higher child or adult GRS. § Percentage difference in change per year, per SD higher child or adult GRS. GRS, genetic risk score; SD, standard deviation; CI, confidence interval. | | | | |
