## Supplemental File 1 - STROBE for "Life stage-specific effects of genetic susceptibility to higher body size on body fat and lean mass: prospective cohort study"

STROBE Statement—checklist of items that should be included in reports of observational studies

|  | Item No. | Recommendation | Page  No. |
| --- | --- | --- | --- |
| **Title and abstract** | 1 | (*a*) Indicate the study’s design with a commonly used term in the title or the abstract | Abstract: Methods |
|  |  | (*b*) Provide in the abstract an informative and balanced summary of what was done and what was found | Abstract: Conclusions |
| Introduction | | | |
| Background/rationale | 2 | Explain the scientific background and rationale for the investigation being reported | Introduction: Paragraph 2 |
| Objectives | 3 | State specific objectives, including any prespecified hypotheses | Introduction: paragraph 2 |
| Methods | | | |
| Study design | 4 | Present key elements of study design early in the paper | Methods: Paragraph 4 |
| Setting | 5 | Describe the setting, locations, and relevant dates, including periods of recruitment, exposure, follow-up, and data collection | Methods: Paragraph 1 |
| Participants | 6 | (*a*) *Cohort study*—Give the eligibility criteria, and the sources and methods of selection of participants. Describe methods of follow-up  *Case-control study*—Give the eligibility criteria, and the sources and methods of case ascertainment and control selection. Give the rationale for the choice of cases and controls  *Cross-sectional study*—Give the eligibility criteria, and the sources and methods of selection of participants | Methods: Paragraph 5 |
|  |  | (*b*) *Cohort study*—For matched studies, give matching criteria and number of exposed and unexposed  *Case-control study*—For matched studies, give matching criteria and the number of controls per case |  |
| Variables | 7 | Clearly define all outcomes, exposures, predictors, potential confounders, and effect modifiers. Give diagnostic criteria, if applicable | Methods: Paragraph 3-4 |
| Data sources/ measurement | 8* | For each variable of interest, give sources of data and details of methods of assessment (measurement). Describe comparability of assessment methods if there is more than one group | Methods: Paragraphs 3-4 |
| Bias | 9 | Describe any efforts to address potential sources of bias |  |
| Study size | 10 | Explain how the study size was arrived at | Methods:  Paragraph 5 |

Continued on next page

| Quantitative variables | 11 | Explain how quantitative variables were handled in the analyses. If applicable, describe which groupings were chosen and why | Methods: Paragraph 6 | To aid interpretation, GRSs were internally standardised by centring around the sample mean and dividing by the SD: (individual GRS minus mean(GRS) divided by SD(GRS). Fat-based DXA measures had skewed distributions on most occasions and were therefore log transformed before modelling took place. |
| --- | --- | --- | --- | --- |
| Statistical methods | 12 | (*a*) Describe all statistical methods, including those used to control for confounding | Methods: Paragraph 5 | all analyses were performed with mutual adjustment for GRSs, i.e., the childhood GRS model was adjusted for the adulthood GRS and vice versa. Multilevel models estimate the mean trajectory of the outcome while accounting for the non-independence (i.e., clustering) of repeated measurements within individuals, change in scale and variance of measures over time, and differences in the number and timing of measurements between individuals (using all available data from all eligible participants under a Missing at Random (MAR) assumption). Linear splines allow knot points to be fit at different ages to derive periods in which change is approximately linear. Knot points were fitted at age 13 and 15 as per previous work, with an additional knot point added at 18 for the extension of trajectories to 25 years. These linear spline multilevel models included two levels: measurement occasion and individual. All DXA models included sex- and age-specific adjustments for height as previously described |
|  |  | (*b*) Describe any methods used to examine subgroups and interactions | Methods: Paragraph 5 |  |
|  |  | (*c*) Explain how missing data were addressed | Methods:  Paragraph 5 | Only complete cases were used as per inclusion criteria |
|  |  | (*d*) *Cohort study*—If applicable, explain how loss to follow-up was addressed  *Case-control study*—If applicable, explain how matching of cases and controls was addressed  *Cross-sectional study*—If applicable, describe analytical methods taking account of sampling strategy | Methods:  Paragraph 5 | using all available data from all eligible participants under a Missing at Random (MAR) assumption |
|  |  | (*e*) Describe any sensitivity analyses | Methods:  Paragraph 5 | Sensitivity analysis models as described above were also run using a a) non-sex specific GRS, b) without mutual adjustment and c) non sex-specific GRS without mutual adjustment. |
| Results | | | | |
| Participants | 13* | (a) Report numbers of individuals at each stage of study—eg numbers potentially eligible, examined for eligibility, confirmed eligible, included in the study, completing follow-up, and analysed | Results: Paragraph 1 | A total of 6926 individuals (3511 females and 3415 males) were included in our analyses. |
|  |  | (b) Give reasons for non-participation at each stage |  | Inclusion criteria covers this |
|  |  | (c) Consider use of a flow diagram | NA |  |
| Descriptive data | 14* | (a) Give characteristics of study participants (eg demographic, clinical, social) and information on exposures and potential confounders | Results: Table 1 |  |
|  |  | (b) Indicate number of participants with missing data for each variable of interest | NA | Inclusion criteria & methodology deals with missing data |
|  |  | (c) *Cohort study*—Summarise follow-up time (eg, average and total amount) | Table 1 | Details age 9 to age 25 DXA values |
| Outcome data | 15* | *Cohort study*—Report numbers of outcome events or summary measures over time | Table 1 | Details body composition measures analysed |
|  |  | *Case-control study—*Report numbers in each exposure category, or summary measures of exposure |  |  |
|  |  | *Cross-sectional study—*Report numbers of outcome events or summary measures |  |  |
| Main results | 16 | (*a*) Give unadjusted estimates and, if applicable, confounder-adjusted estimates and their precision (eg, 95% confidence interval). Make clear which confounders were adjusted for and why they were included | Results: Paragraph 3 | A 1 SD higher childhood GRS or adult GRS was associated with 8.8% (95% Confidence interval (CI) 6.3, 11.3) and 6.1% (95% CI 3.7, 8.6) higher total fat mass at 9y, |
|  |  | (*b*) Report category boundaries when continuous variables were categorized | NA |  |
|  |  | (*c*) If relevant, consider translating estimates of relative risk into absolute risk for a meaningful time period | NA |  |

Continued on next page

| Other analyses | 17 | Report other analyses done—eg analyses of subgroups and interactions, and sensitivity analyses | Results: Paragraph 5 | Unadjusted findings (from models that did not adjust for the other GRS) were broadly similar to adjusted findings among males and females (**Supplementary Tables 2-3**). Mutually adjusted models using sex-combined GRS were also used |
| --- | --- | --- | --- | --- |
| Discussion | | | | |
| Key results | 18 | Summarise key results with reference to study objectives | Discussion:  Paragraph 1 | Our findings show that genetic variants for body size are more strongly associated with adiposity than lean mass from childhood to early adulthood. They also demonstrate that childhood variants are more strongly associated with adiposity in females than adulthood variants until early adulthood whereas childhood and adulthood variants are similarly associated with adiposity across early life in males. Our findings may inform selection of instruments for life stage- specific adiposity in future MR studies |
| Limitations | 19 | Discuss limitations of the study, taking into account sources of potential bias or imprecision. Discuss both direction and magnitude of any potential bias | Discussion: Paragraph 4 | The limitations of this study include the restriction of analyses to participants of white ethnicity (given the European sample from which genetic variants were discovered) and therefore generalisability of our findings other ethnic groups |
| Interpretation | 20 | Give a cautious overall interpretation of results considering objectives, limitations, multiplicity of analyses, results from similar studies, and other relevant evidence | Discussion: Paragraph 3 | Our results suggest that genetic variants for body size are more strongly associated with body fat than lean mass across early life in both sexes |
| Generalisability | 21 | Discuss the generalisability (external validity) of the study results | Conclusion: Paragraph 1 | Findings may inform selection of instruments for life stage specific adiposity in future Mendelian randomization studies |
| Other information | |  | | |
| Funding | 22 | Give the source of funding and the role of the funders for the present study and, if applicable, for the original study on which the present article is based | Funding Subheading | SW is supported by Cancer Research UK [grant number C18281/A30905]. TGR, GDS, and JAB work in a unit funded by the UK MRC (MC_UU_00011/1) and the University of Bristol. LMOK is supported by a Health Research Board (HRB) of Ireland Emerging Investigator Award (EIA-FA-2019-007 SCaRLeT). The UK Medical Research Council and Wellcome (Grant ref: 217065/Z/19/Z) and the University of Bristol provide core support for ALSPAC. A comprehensive list of grants funding is available on the ALSPAC website (<http://www.bristol.ac.uk/alspac/external/documents/grant-acknowledgements.pdf>) and GWAS data was generated by Sample Logistics and Genotyping Facilities at Wellcome Sanger Institute and LabCorp (Laboratory Corporation of America) using support from 23andMe. This publication is the work of the authors and SW is the guarantor for its contents. The funders had no role in study design, data collection and analysis, decision to publish, or preparation of the manuscript. |

*Give information separately for cases and controls in case-control studies and, if applicable, for exposed and unexposed groups in cohort and cross-sectional studies.

**Note:** An Explanation and Elaboration article discusses each checklist item and gives methodological background and published examples of transparent reporting. The STROBE checklist is best used in conjunction with this article (freely available on the Web sites of PLoS Medicine at http://www.plosmedicine.org/, Annals of Internal Medicine at http://www.annals.org/, and Epidemiology at http://www.epidem.com/). Information on the STROBE Initiative is available at www.strobe-statement.org.
